# Impact of nucleos(t)ide analogues on the risk of hepatocellular carcinoma in chronic hepatitis B patients: A time-dependent Cox regression analysis

**DOI:** 10.1101/2023.09.27.23296247

**Authors:** Makoto Moriyama, Ryosuke Tateishi, Mizuki Nishibatake Kinoshita, Tsuyoshi Fukumoto, Tomoharu Yamada, Taijiro Wake, Ryo Nakagomi, Takuma Nakatsuka, Tatsuya Minami, Masaya Sato, Mitsuhiro Fujishiro, Kazuhiko Koike

## Abstract

**Background and Aims:** The preventive effect of nucleos(t)ide analog (NA) use on HCC development in patients with chronic hepatitis B (CHB) is controversial due to the difficulty of conducting randomized controlled trials.

**Approach and Results:** In this single-center, retrospective study, NA-naïve CHB patients without a history of HCC were enrolled and followed-up from the first visit on or after January 2000 to December 2020. Patients were categorized into the NA group, including those who started NA after study enrollment, and the non-NA group, including patients to whom NA was never administered during the follow-up period. After propensity score matching (PSM) to balance the confounding factors, we applied a multivariable time-dependent Cox proportional regression analysis with the initiation of NA as a time-dependent covariate. We further performed a subgroup analysis according to the presence or absence of cirrhosis. The baseline characteristics of 212 pairs of patients retrieved by PSM were comparable. During the mean follow-up of 12.9 and 6.8 years in the NA and non-NA groups, respectively, 25 and 28 patients developed HCC, respectively. Multivariable analysis with time-dependent covariates showed that NA did not affect HCC risk (HR, 0.68; 95% CI, 0.36–1.31; *p* = 0.25) after adjusting for other risk factors, including age, sex, and HBV viral load. Subgroup analysis showed that NA use significantly reduced the risk of HCC in cirrhotic patients (HR, 0.26; 95% CI, 0.08–0.85; *p* = 0.03).

**Conclusions:** The preventive effect of NA on hepatocarcinogenesis may be limited to cirrhotic patients.

## INTRODUCTION

Approximately 350 million people worldwide are chronically infected with HBV.[1–3] Chronic hepatitis B (CHB) is a leading cause of liver-related adverse events, including liver cirrhosis and HCC. The annual incidence of HBV-related HCC varies according to risk factors, from <0.1% in health carriers to 2% to 5% in cirrhosis.[4] Previous studies have reported that baseline HBV DNA load is a significant risk factor for HCC.[5, 6]

Administration of nucleos(t)ide analog (NA) in the case of CHB decreases the viral load and suppresses liver fibrosis progression.[7, 8] This suggests that the administration of NA in the case of CHB reduces HCC incidence. One randomized controlled trial supported the concept comparing lamivudine administration to placebo in patients with cirrhosis and CHB, where HCC incidence assessed as a part of the composite outcome, was less frequent in the lamivudine group with a marginal significance (*p* = 0.047).[9] The unsustainable effect of lamivudine partially explains the reason for the marginal outcome.[10, 11] Therefore, more potent NAs with lasting effects, including entecavir and tenofovir, may exert a more pronounced preventive effect.[12–14] However, as the beneficial effect of NA on CHB, particularly reduction in risk of hepatic decompensation, has become apparent, it is no longer ethically feasible to conduct randomized controlled trials to evaluate the efficacy of NA on hepatocarcinogenesis.

Several observational studies have shown the risk reduction of NA on HCC.[15–20] However, comparing treated and untreated groups reveal the following issues. First, if the observation period in the treated group started at the beginning of the treatment and at the patient’s first visit to the clinic in the untreated group, the latter has a substantially longer observation period. Second, if the observation began at the first visit to the clinic, and patients were divided into treated and untreated groups after enrollment, there is an immortal time bias: patients in the treated group will not experience the outcome during part of the follow-up period (Supplementary Figure 1). [21] Cox proportional hazard model with time-dependent covariates or landmark analysis is recommended to overcome these issues.[22, 23] Here, we aimed to evaluate the effect of NA on hepatocarcinogenesis in CHB patients using the aforementioned statistical methods.

## PATIENTS AND METHODS

### Study protocol

This retrospective study was conducted according to the ethical guidelines for epidemiological research established by the Japanese Ministry of Education, Culture, Sports, Science and Technology and Ministry of Health, Labour, and Welfare. The study design was included in a comprehensive protocol of retrospective studies at the Department of Gastroenterology, the University of Tokyo Hospital, and approved by the University of Tokyo Medical Research Center Ethics Committee (approval no. 2058).

Informed consent was waived because of the retrospective design. The following statements were posted at a website (http://gastro.m.u-tokyo.ac.jp/patient/clinicalresearch.html) and participants who do not agree to the use of their clinical data can claim deletion of them.

### Data collection

We collected data from CHB patients diagnosed at our department since 1985, which was stored in a designated computerized database. We retrieved clinical data from the first visit after January 1, 2000 to December 31, 2020. These data included baseline characteristics: age, sex, presence of cirrhosis, and laboratory data, including total bilirubin, albumin, aspartate aminotransferase (AST), alanine aminotransferase (ALT), platelet count, HBeAg positivity, and HBV DNA load. The viral load was converted to IU/mL using the conversion formula described in Supplementary Table 1, as the measurement methods for HBV DNA changed during the study period. Cirrhosis was diagnosed based on clinical findings, laboratory data, imaging findings, and liver stiffness measured by transient elastography or liver biopsy. The database also stored the treatment regimen, the date of treatment initiation, and the response regarding NA use.

### Patient enrollment

Inclusion criteria were as follows: (i) chronic infection with HBV, defined as being positive for hepatitis B surface antigen (HBsAg) for at least 6 months, and (ii) patients over 18 years of age. Exclusion criteria were as follows: (i) coinfection with chronic hepatitis C, (ii) history of HCC before enrollment, (iii) second opinion or referral cases without follow-up, (iv) those who received NA before study enrollment, and (v) those whose HBV DNA load was not measured at enrollment. The patients were divided into the NA group, which included patients who started NA after enrollment in the study, and the non-NA group, which included patients in whom NA was never administered during the follow-up period. According to the Japanese clinical practice guidelines for CHB [24], patients were recommended to receive NA with serum HBV DNA level >2 000 IU/mL and elevated ALT level (>31U/L), or with advanced fibrosis. However, some patients refused to start NA treatment despite meeting the criteria.

### Definition of viral response

Sustained virological response (SR) during NA treatment is defined as undetectable HBV DNA by a sensitive polymerase chain reaction (PCR) assay with a limit of detection according to the clinical practice guidelines for CHB[24, 25]. The lower limit of detection sensitivity differs depending on the time of measurement and testing methods (Supplementary Table 1). Primary non-response was defined as less than one log_10_ IU/mL reduction of HBV DNA after 3 months of therapy. Virological breakthrough is defined as a confirmed increase in HBV DNA level of more than 1 log_10_ IU/mL compared to the lowest value HBV DNA level on-therapy.

### Follow-up and diagnosis of HCC

Patients were followed-up at the outpatient clinic with blood tests, including serum alpha-fetoprotein (AFP), *lens culinaris* agglutinin-reactive fraction of alpha-fetoprotein (AFP-L3), and des-gamma-carboxy-prothrombin (DCP) levels and ultrasonography at every 6 months according to Japanese clinical practice guidelines for HCC.[26] Contrast-enhanced CT or MRI were performed when tumors were detected on ultrasonography or when tumor markers were elevated, suggestive of HCC. Patients were followed-up until any confirmed HCC diagnosis or the last visit before December 31, 2021. Data of patients who died from any cause without HCC diagnosis or who had a liver transplant were censored.

The study endpoint was the development of HCC. Considering hyper-attenuation in the arterial phase and washout in the late phase as definite signs of HCC, a diagnosis of HCC was made by dynamic CT or MRI.[27] When the imaging diagnosis was indeterminate, we confirmed HCC pathologically by ultrasound-guided tumor biopsy.

### Statistical analysis

Data are expressed as medians with 25^th^ to 75^th^ percentiles unless otherwise indicated. Numbers and percentages were used for qualitative variables. The categorical variables were compared with χ^2^ tests, ordinal variables were compared with the Cochran– Armitage test, and continuous variables were compared with unpaired Student’s *t*-tests. To normalize the two groups (NA and non-NA groups), propensity score matching (PSM), including age, sex, presence of cirrhosis, total bilirubin, albumin, AST, ALT, platelet count, HBeAg positivity, and HBV DNA >2 000 IU/mL at baseline, was performed. In PSM analysis, we used logistic regression to estimate the probability of a patient to start NA treatment and generated a propensity score for each patient. Caliper matching on the propensity score was performed, and pairs were matched within a range of 0.2 of the standard deviation of the logit of the propensity score. The cumulative incidence of HCC was assessed with the Kaplan–Meier method. For the estimation of cumulative incidence of HCC with time-dependent grouping, we used the Simon and Makuch method, which was a computational method to graphically represent survival curves for time-dependent covariates.[28] We also estimated the prognosis after HCC development with the Kaplan–Meier method.

We performed univariable and multivariable Cox proportional regression analyses using time-fixed and time-dependent covariates in the PSM cohort. Exposure to NA in addition to albumin, ALT, platelet count, and HBV DNA load were treated as time-dependent covariates. We further performed a subgroup analysis according to the presence or absence of cirrhosis. We also performed 1-year and 2-year landmark analyses in the PSM cohort to mitigate the immortal time bias, where patients were stratified according to NA use prior to the corresponding time points.

Statistical analyses were performed using the R software (Ver. 4.1.3; R Development Core Team, Vienna, Austria). All tests were two-tailed. *p*-values < 0.05 were considered statistically significant.

## RESULTS

### Patient profiles

Among the 1612 patients with CHB identified using a database search, 884 fulfilled the enrollment criteria (Figure 1). A total of 274 patients started receiving NA after enrollment (NA group) and 610 patients did not receive NA (non-NA group). The median interval from the study enrollment to the initiation of NA therapy was 2.19 (0.35-5.66) years, and the median duration of NA administration was 7.83 (3.80–12.0) years. The baseline characteristics of the entire cohort and the matched cohort are shown in Table 1. Significant differences were found in terms of sex, presence of cirrhosis, total bilirubin levels, albumin levels, AST, ALT, platelet count, the proportion of those with HBeAg positivity, and HBV DNA>2 000 IU/mL. The NA group included a higher proportion of patients with HBeAg, presence of cirrhosis, and high viral HBV load. The baseline characteristics of the 212 pairs of matched patients from the two study groups were comparable.

**Figure 1.**
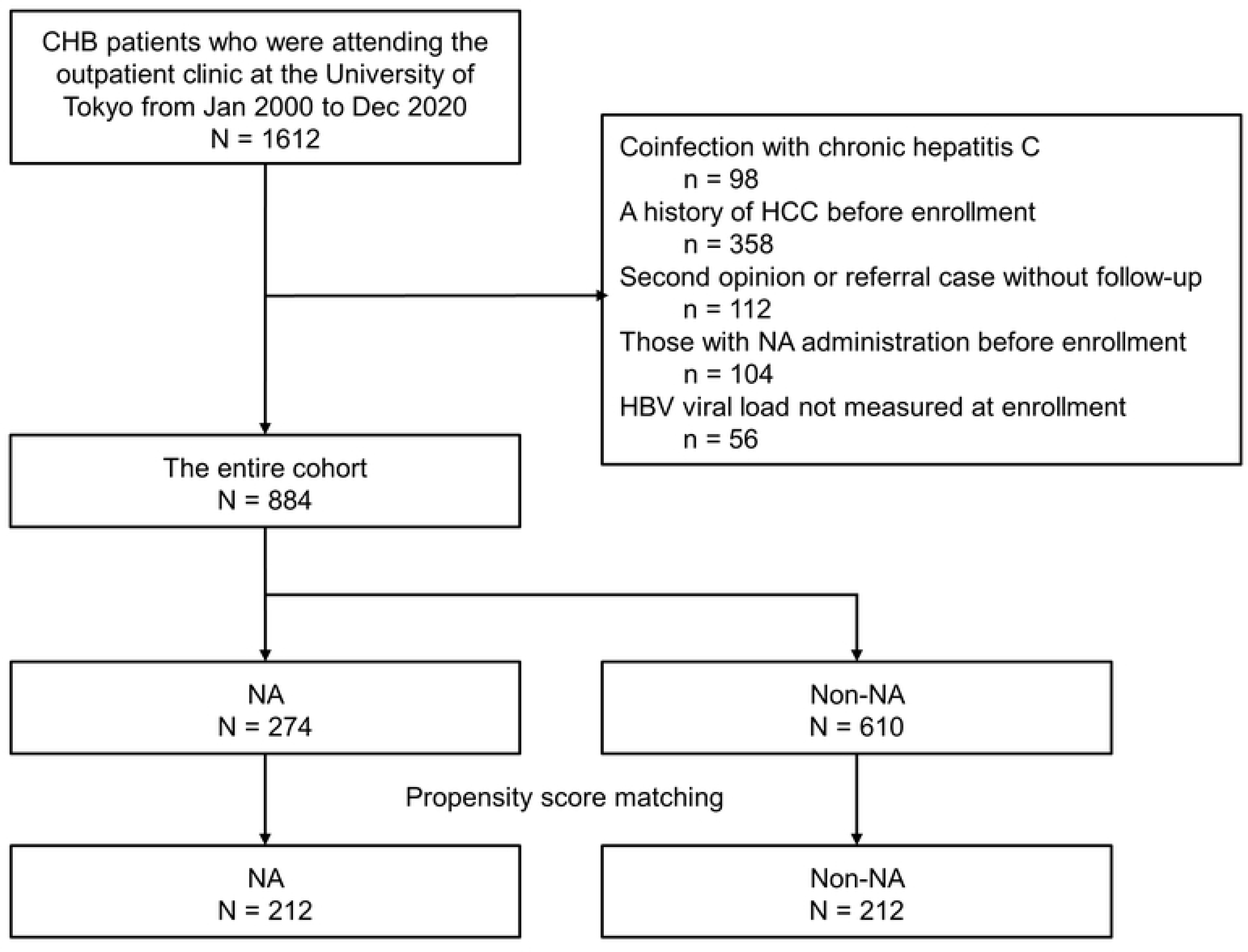
Patient enrollment flow.

**Table 1.**
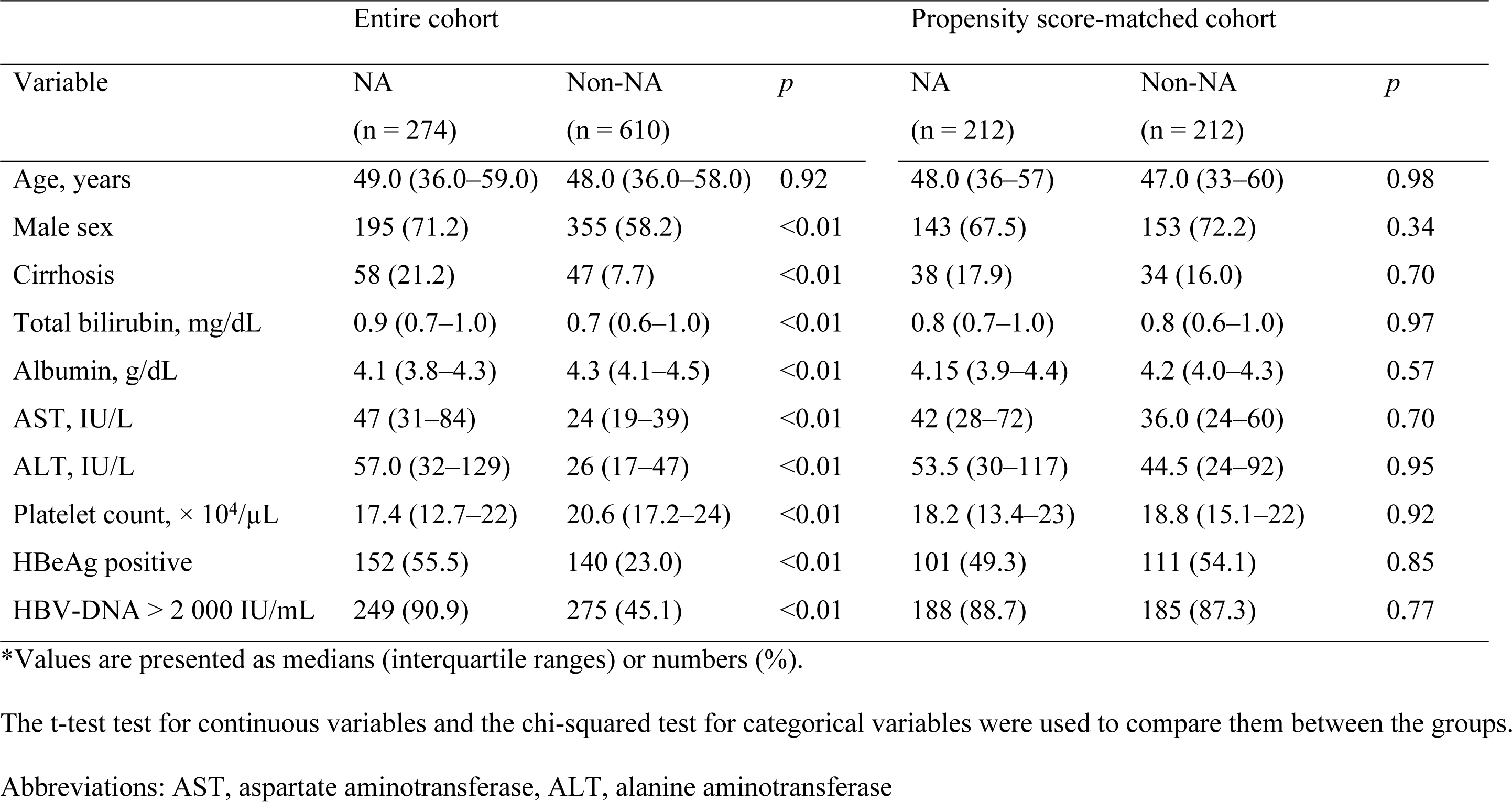
Baseline characteristics of the entire cohort (N = 884) and Propensity score-matched cohort (N = 424) *.

### Nucleic acid analogs and viral response

Among 274 patients in the NA group, 74 patients received lamivudine (LAM), 182 received entecavir (ETV), 5 received tenofovir disoproxil fumarate (TDF), and 13 received tenofovir alafenamide (TAF) (Supplementary Figure 2A,2B). As adefovir (ADV) monotherapy is not covered by health insurance in Japan, none of the patients were treated with ADV monotherapy. Treatment response could be evaluated in 72 of 74 patients who started NA treatment with LAM. Among them, primary non-response was observed in 1 who achieved SR with ADV add-on and viral breakthrough was observed in 31: 24 of whom SR was achieved with ADV add-on, 3 with a switch to ETV, and 1 with a switch to TAF. The remaining 40 patients achieved SR with LAM, with 22 and 4 switching to ETV and TAF, respectively, by the end of observation. The viral breakthrough was observed in 3 patients treated with ETV, of whom 2 achieved SR with the addition of TAF and one patient dropped out of follow-up. All patients with TDF and TAF achieved SR. In total, 271 patients achieved SR by the end of the observation period.

### HCC development

In the PSM cohort, during the mean follow-up of 12.9 and 6.8 years in the NA and non-NA groups, respectively, 25 patients of the NA group and 28 of the non-NA group developed HCC. The cumulative incidence rates of HCC development by Kaplan– Meier analysis at 5 and 10 years were 6.0%, 12.7%, respectively (Figure 2A). The survival curves determined for NA use and non-use (time-dependent covariate) using the Simon and Makuch method are shown in Figure 2B.[28]

**Figure 2A.**
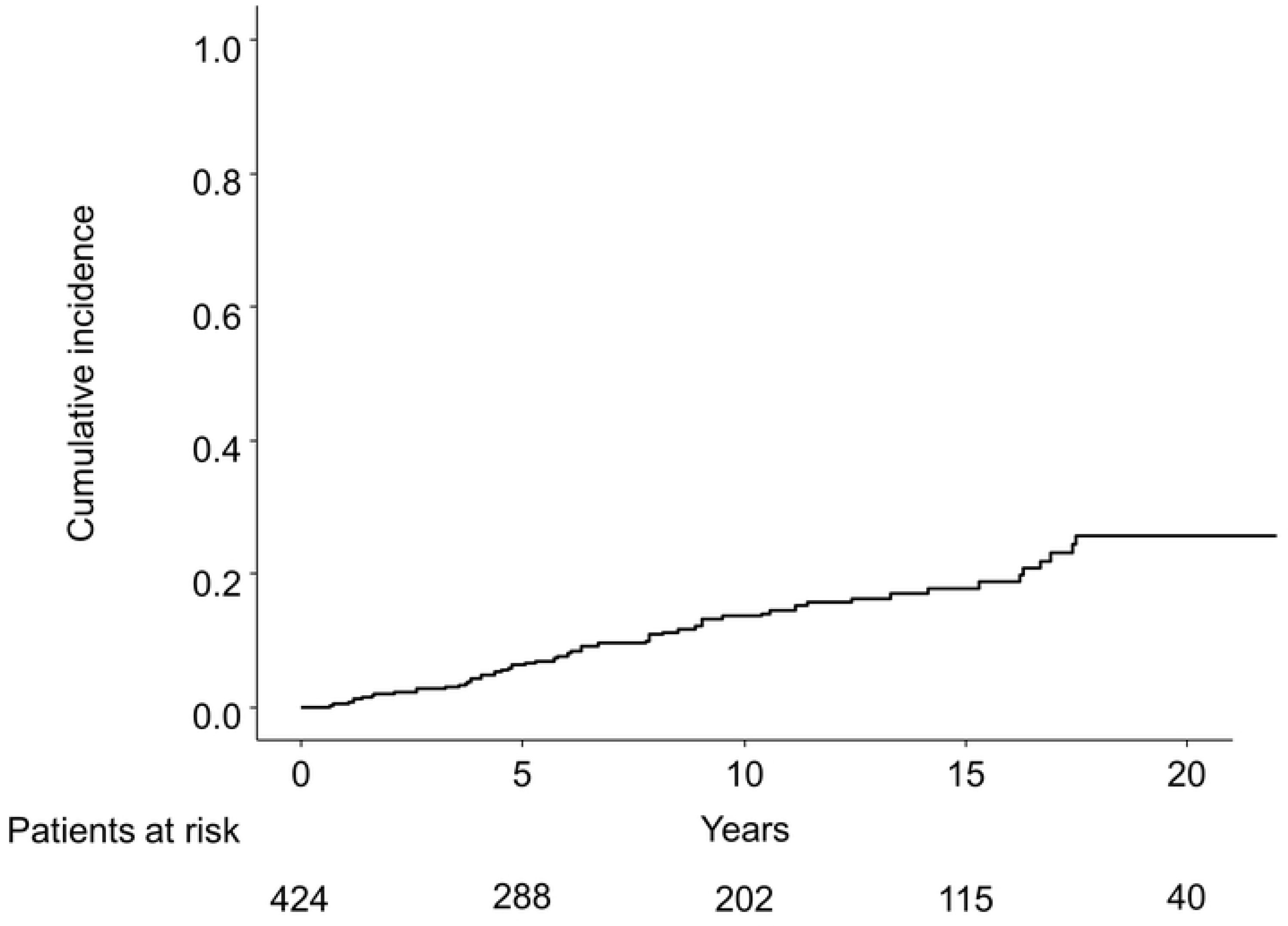
Kaplan–Meier estimate of cumulative incidence of HCC in the PSM cohort (N = 424). HCC, hepatocellular carcinoma; PSM, propensity score matching

**Figure 2B.**
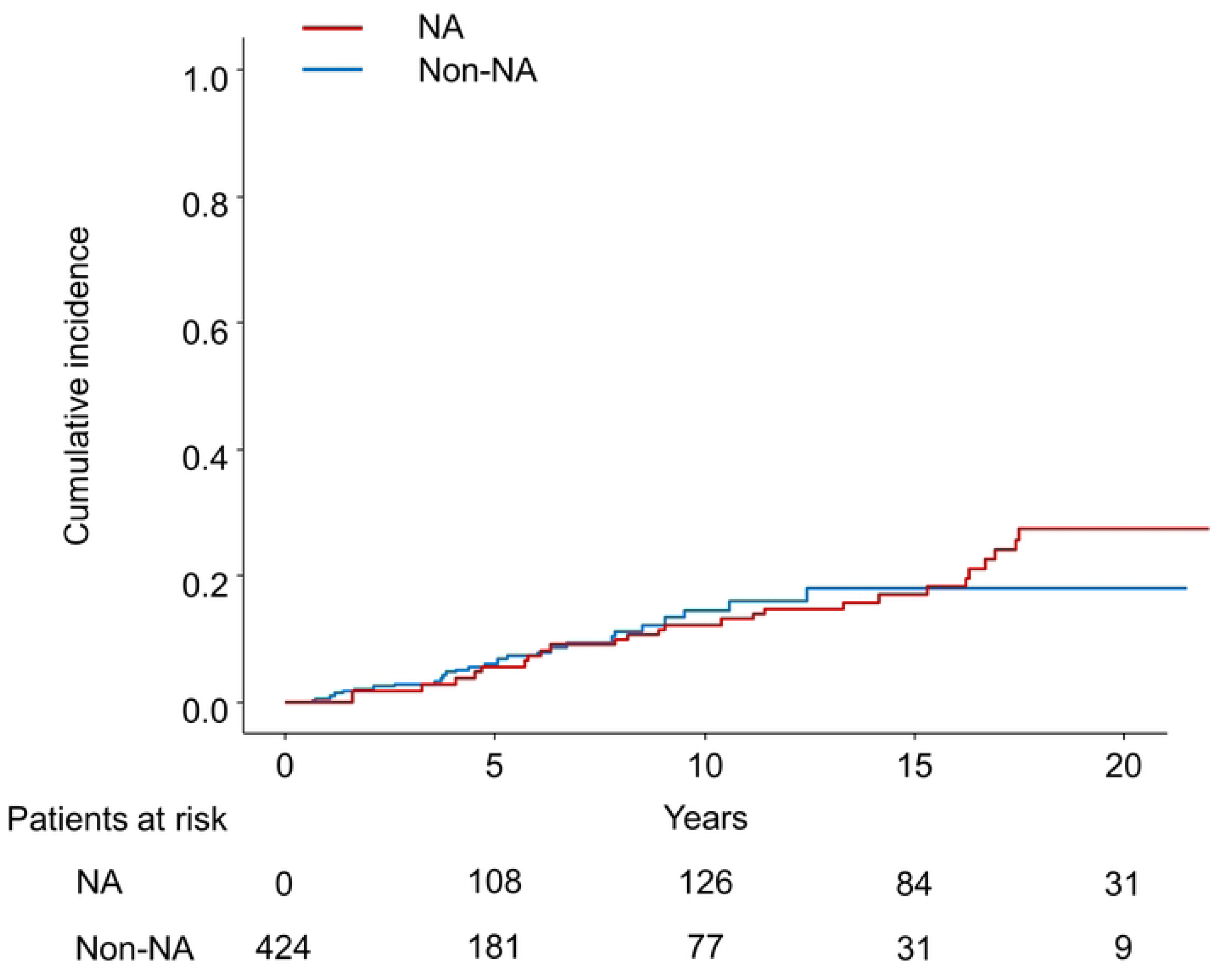
The cumulative incidence of HCC with time-dependent grouping plotted by the Simon and Makuch method. HCC, hepatocellular carcinoma; NA, nucleos(t)ide analog

The characteristics of HCC are shown in Table 2. HCC was diagnosed at an earlier stage, i.e., with smaller size and fewer nodules, in the NA group than in the non-NA group. AST and ALT at the diagnosis of HCC were significantly lower in the NA group than in the non-NA group.

**Table 2.** Characteristics of HCC patients at diagnosis in the propensity score- matched cohort (N = 53)*

| Variable | NA<br>(n = 25) | Non-NA<br>(n = 28) | *p* |
| --- | --- | --- | --- |
| Tumor size, mm | 20 (16–25) | 22 (20.5–30.0) | 0.03 |
| Number of nodules |  |  | 0.09 |
| Solitary | 23 (92.0%) | 19 (67.9%) |  |
| 2–3 | 1 (4%) | 8 (28.6%) |  |
| >3 | 1 (4%) | 1 (3.6%) |  |
| AFP >15 ng/mL | 3 (12%) | 12 (42.9%) | 0.02 |
| AFP-L3 >10% | 1 (4%) | 7 (25%) | 0.07 |
| DCP >100 mAU/mL | 6 (24%) | 5 (17.9%) | 0.89 |
| Total bilirubin (mg/dL) | 0.8 (0.6–1) | 0.8 (0.6–1.0) | 0.57 |
| Albumin (g/dL) | 4.0 (3.9–4.3) | 4.0 (3.6–4.2) | 0.21 |
| AST (IU/L) | 28 (19–31) | 41 (28.5–47) | 0.01 |
| ALT (IU/L) | 19 (14–27) | 38 (22–51) | 0.01 |
| Platelet count ($\times 10^4/\mu\text{L}$) | 17 (13–20) | 12.8 (10.5–18) | 0.19 |

\*Values are presented as medians (interquartile ranges) or numbers (%).
The t-test for continuous variables, the chi-squared test for categorical variables, and the Cochran–Armitage test for ordinal variables were used to compare the groups.
Abbreviations: AFP, alpha-fetoprotein, AFP-L3, lens culinaris agglutinin-reactive fraction of AFP, DCP, des-gamma-carboxy prothrombin, AST, aspartate aminotransferase, ALT, alanine aminotransferase

### Prognosis after HCC development

Among 53 patients who developed HCC, 21 (5 in the NA group and 16 in the non-NA group) died before the end of the observation period. The median survival time after HCC development was 16.5 years. The survival curves after HCC development according to the NA use at the diagnosis were shown in Supplementary Figure 3. The log-rank test showed no significant difference between the two groups (*p* = 0.3).

### Univariable and multivariable time-dependent Cox regression analysis after PSM

The univariable analysis showed that the following factors were significantly associated with HCC development: age, presence of cirrhosis, albumin level, AST>40 IU/mL, and platelet count. In the univariable analysis, the HR of NA use was 1.10 (95% CI, 0.61– 1.98; *p* = 0.75). In the multivariable analysis, NA use did not significantly decrease the risk of HCC (HR, 0.68; 95% CI, 0.36–1.31; *p* = 0.25) (Table 3).

**Table 3.** Univariable and multivariable analyses of hepatocarcinogenesis in CHB patients (Propensity Score-Matched cohort)

| Variable | Univariable |  | Multivariable |  |
| --- | --- | --- | --- | --- |
|  | HR (95% CI) | *p* | HR (95% CI) | *p* |
| Age per 1 year* | 1.05 (1.03–1.07) | <0.01 | 1.05 (1.02–1.08) | <0.01 |
| Female sex* | 0.87 (0.47–1.61) | 0.67 | 0.68 (0.37–1.27) | 0.36 |
| Cirrhosis* | 5.27 (3.06–9.08) | <0.01 | 4.10 (2.25–7.49) | <0.01 |
| Total bilirubin (mg/dL)<sup>†</sup> | 1.04 (0.90–1.20) | 0.64 |  |  |
| ALB per 1 g/dL<sup>†</sup> | 0.42 (0.29–0.61) | <0.01 | 0.77 (0.48–1.24) | 0.28 |
| AST>40 IU/mL<sup>†</sup> | 2.94 (1.54–5.61) | <0.01 |  |  |
| ALT>40 IU/mL<sup>†</sup> | 1.81 (0.96–3.38) | 0.07 | 1.74 (0.89–3.40) | 0.11 |
| Platelet count ($\times 10^4/\mu\text{L}$)<sup>†</sup> | 0.91 (0.88–0.95) | <0.01 | | |
| HBeAg positive* | 0.88 (0.51–1.52) | 0.65 | 1.20 (0.67–2.14) | 0.54 |
| HBV-DNA >2 000 IU/mL<sup>†</sup> | 1.47 (0.58–3.68) | 0.42 | 1.49 (0.58–3.84) | 0.41 |
| NA use<sup>†</sup> | 1.10 (0.61–1.98) | 0.75 | 0.68 (0.36–1.31) | 0.25 |

\* These variables were analyzed as time-fixed covariates.
<sup>†</sup> These variables were analyzed as time-dependent covariates.
Abbreviations: AST, aspartate aminotransferase; ALT, alanine aminotransferase; NA, nucleos(t)ide analog

### Subgroup analysis

The PSM cohort was divided into 72 cirrhotic and 352 noncirrhotic patients, each analyzed according to the presence or absence of cirrhosis. HCC developed in 23 (10 in the NA group and 13 in the non-NA group) and 32 (17 in the NA group and 15 in the non-NA group) of cirrhotic and noncirrhotic patients, respectively. The survival curves determined for NA use and non-use using the Simon and Makuch method are shown in Figures 3A and 3B. There was a significant difference between the use and non-use of NA in cirrhotic patients (*p =* 0.04), but not in non-cirrhotic patients (*p* = 0.32). The results of univariable and multivariable Cox proportional regression analyses using time-fixed and time-dependent covariates are shown in Tables 4 and 5. In multivariable Cox proportional hazards analysis adjusted for time-fixed and time-dependent covariates, NA use significantly reduced the risk of HCC (HR, 0.26; 95% CI, 0.08– 0.85; p=0.03) only in cirrhotic patients.

**Figure 3A.**
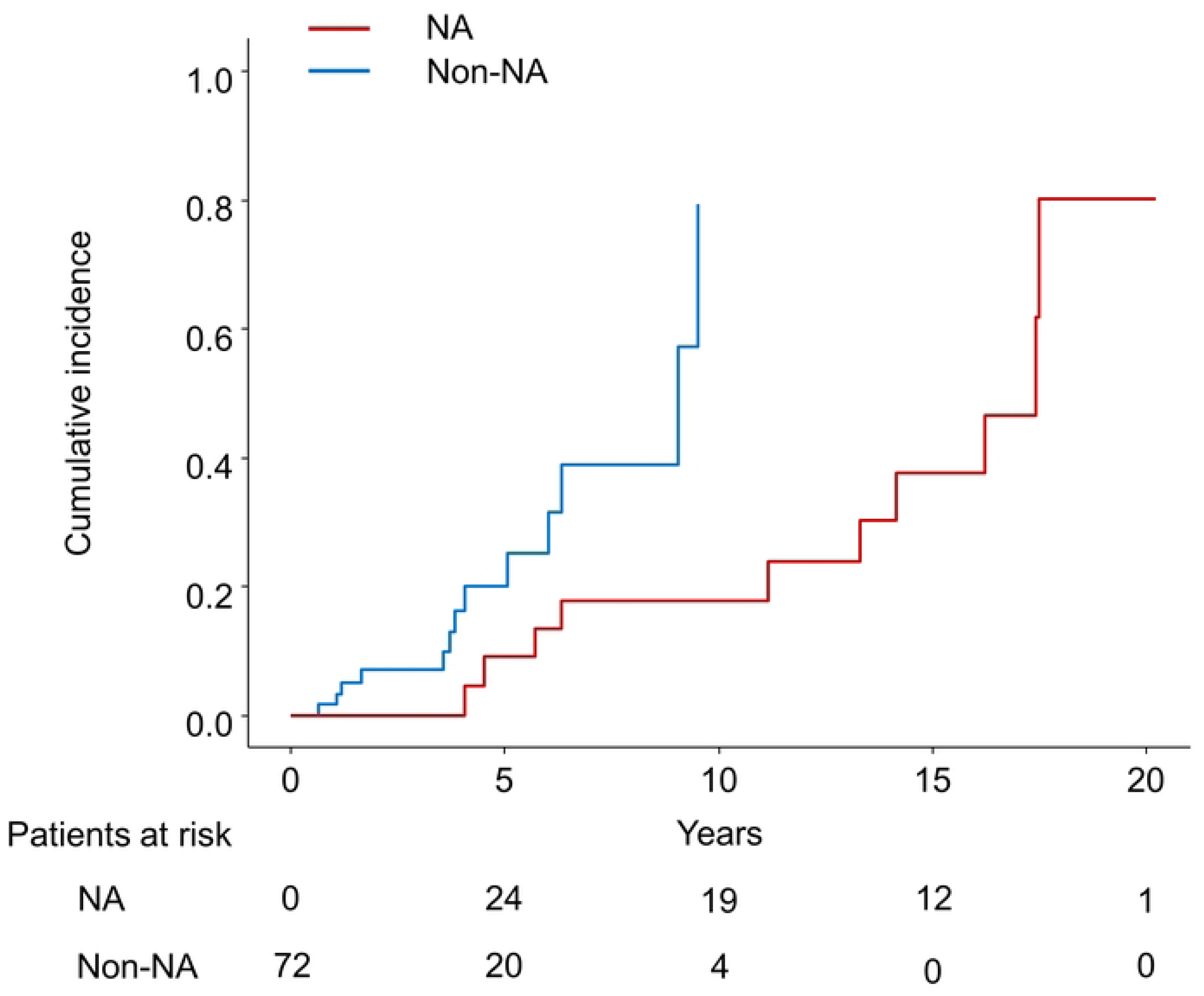
The cumulative incidence of HCC in cirrhotic patients with time-dependent grouping plotted by the Simon and Makuch method. HCC, hepatocellular carcinoma; NA: nucleos(t)ide analog

**Figure 3B.**
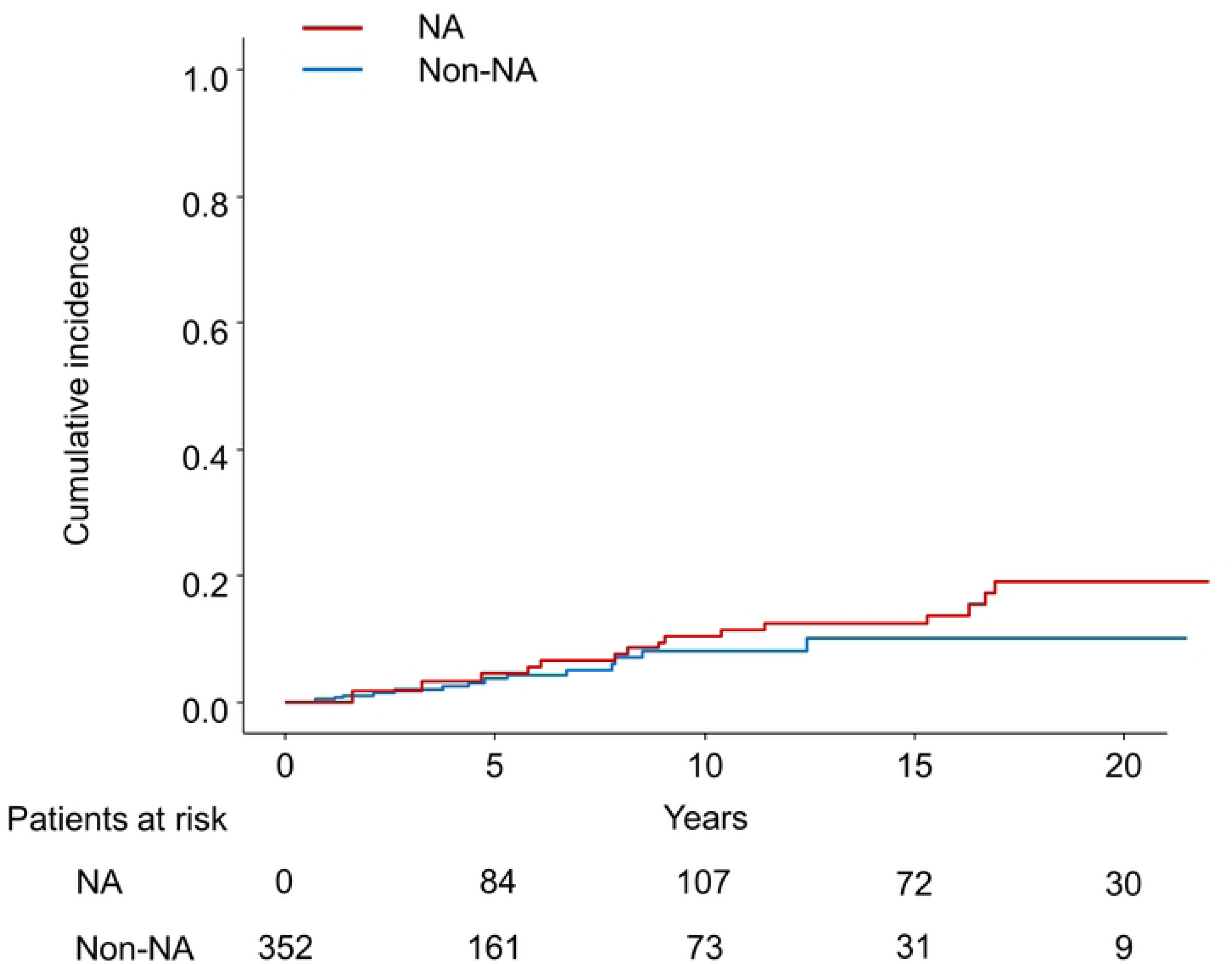
The cumulative incidence of HCC in noncirrhotic patients with time- dependent grouping plotted by the Simon and Makuch method. HCC, hepatocellular carcinoma; NA, nucleos(t)ide analog

**Table 4.**
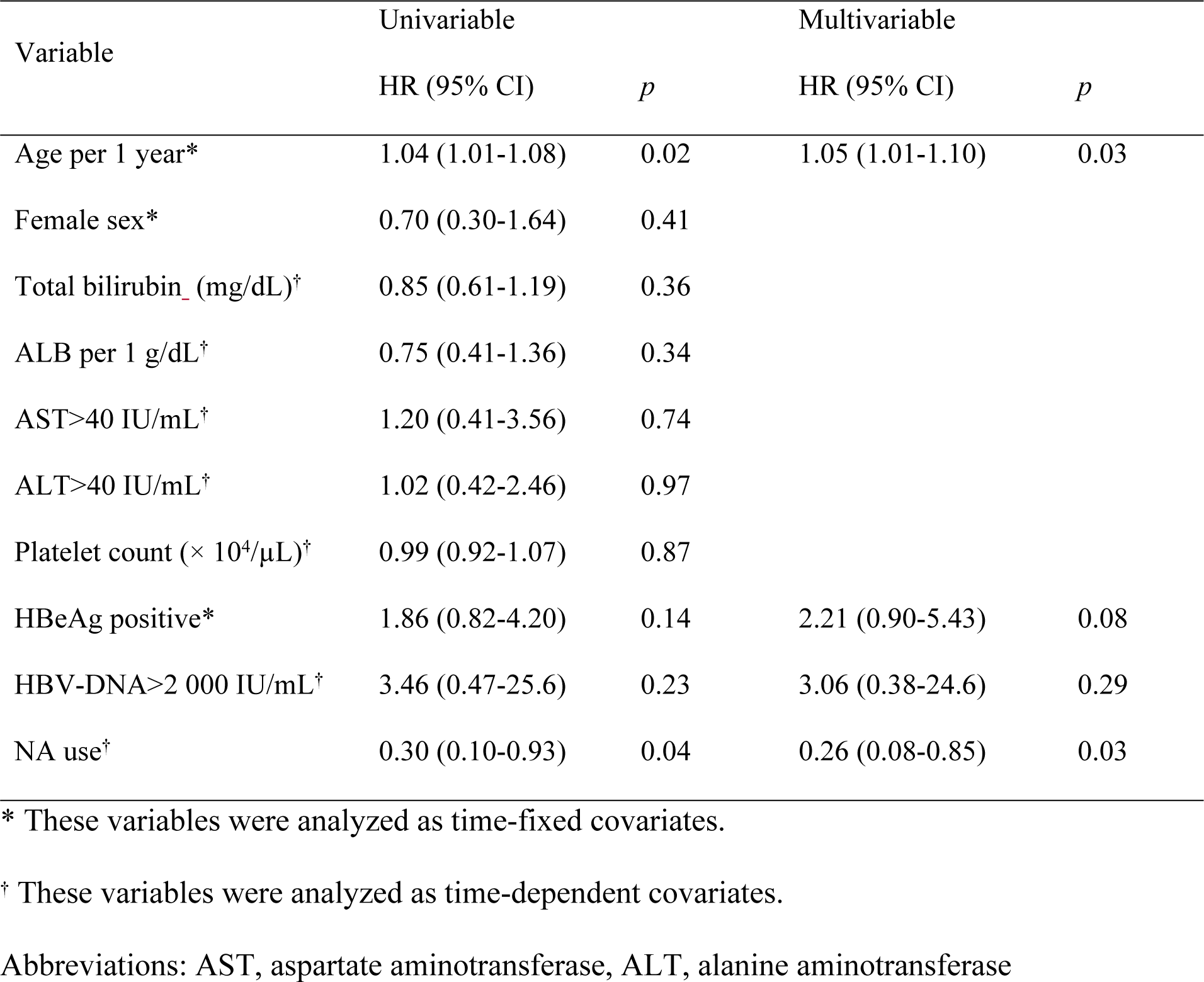
Univariable and multivariable analyses of hepatocarcinogenesis: A.

| Variable | Univariable |  | Multivariable |  |
| --- | --- | --- | --- | --- |
|  | HR (95% CI) | *p* | HR (95% CI) | *p* |
| Age per 1 year* | 1.04 (1.01-1.08) | 0.02 | 1.05 (1.01-1.10) | 0.03 |
| Female sex* | 0.70 (0.30-1.64) | 0.41 |  |  |
| Total bilirubin (mg/dL)<sup>†</sup> | 0.85 (0.61-1.19) | 0.36 |  |  |
| ALB per 1 g/dL<sup>†</sup> | 0.75 (0.41-1.36) | 0.34 |  |  |
| AST>40 IU/mL<sup>†</sup> | 1.20 (0.41-3.56) | 0.74 |  |  |
| ALT>40 IU/mL<sup>†</sup> | 1.02 (0.42-2.46) | 0.97 |  |  |
| Platelet count ($\times 10^4/\mu\text{L}$)<sup>†</sup> | 0.99 (0.92-1.07) | 0.87 | | |
| HBeAg positive* | 1.86 (0.82-4.20) | 0.14 | 2.21 (0.90-5.43) | 0.08 |
| HBV-DNA>2 000 IU/mL<sup>†</sup> | 3.46 (0.47-25.6) | 0.23 | 3.06 (0.38-24.6) | 0.29 |
| NA use<sup>†</sup> | 0.30 (0.10-0.93) | 0.04 | 0.26 (0.08-0.85) | 0.03 |

\* These variables were analyzed as time-fixed covariates.
<sup>†</sup> These variables were analyzed as time-dependent covariates.
Abbreviations: AST, aspartate aminotransferase, ALT, alanine aminotransferase

**Table 5.**
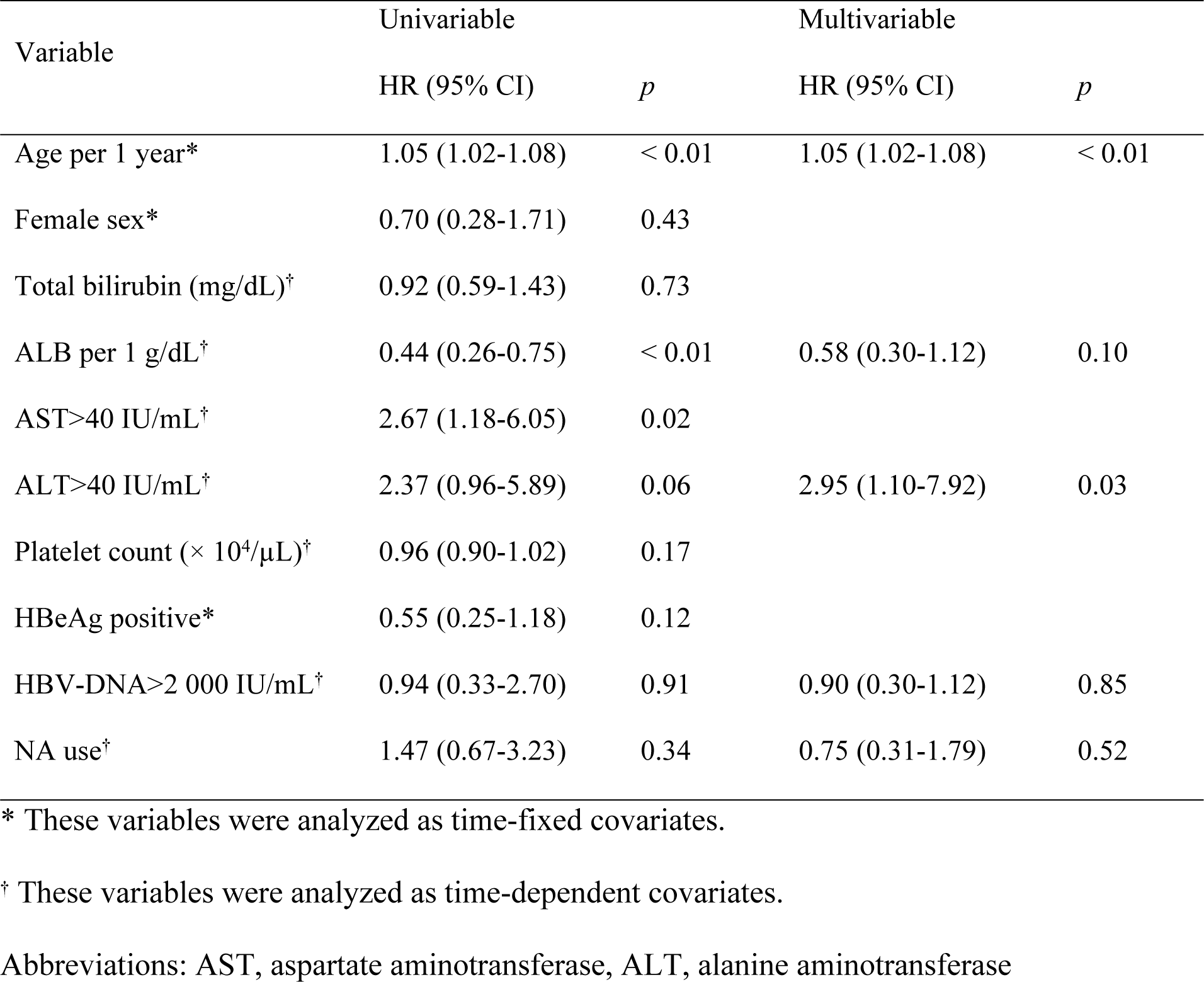
Univariable and multivariable analyses of hepatocarcinogenesis: A subgroup analysis in noncirrhotic patients.

| Variable | Univariable |  | Multivariable |  |
| --- | --- | --- | --- | --- |
|  | HR (95% CI) | *p* | HR (95% CI) | *p* |
| Age per 1 year* | 1.05 (1.02-1.08) | < 0.01 | 1.05 (1.02-1.08) | < 0.01 |
| Female sex* | 0.70 (0.28-1.71) | 0.43 |  |  |
| Total bilirubin (mg/dL)<sup>†</sup> | 0.92 (0.59-1.43) | 0.73 |  |  |
| ALB per 1 g/dL<sup>†</sup> | 0.44 (0.26-0.75) | < 0.01 | 0.58 (0.30-1.12) | 0.10 |
| AST>40 IU/mL<sup>†</sup> | 2.67 (1.18-6.05) | 0.02 |  |  |
| ALT>40 IU/mL<sup>†</sup> | 2.37 (0.96-5.89) | 0.06 | 2.95 (1.10-7.92) | 0.03 |
| Platelet count ($\times 10^4/\mu\text{L}$)<sup>†</sup> | 0.96 (0.90-1.02) | 0.17 | | |
| HBeAg positive* | 0.55 (0.25-1.18) | 0.12 |  |  |
| HBV-DNA>2 000 IU/mL<sup>†</sup> | 0.94 (0.33-2.70) | 0.91 | 0.90 (0.30-1.12) | 0.85 |
| NA use<sup>†</sup> | 1.47 (0.67-3.23) | 0.34 | 0.75 (0.31-1.79) | 0.52 |

\* These variables were analyzed as time-fixed covariates.
<sup>†</sup> These variables were analyzed as time-dependent covariates.
Abbreviations: AST, aspartate aminotransferase, ALT, alanine aminotransferase

### Landmark analysis

We also performed 1-year and 2-year landmark analyses adjusting for age, sex, presence of cirrhosis, albumin, ALT, and HBV DNA load in the multivariable Cox proportional hazard model. Among the 395 patients who were under observation beyond 1 year after enrollment, 53 patients started NA treatment within 1 year. In the cohort, 51 patients developed HCC (4 patients with NAs and 47 without NAs) (Figure 4A). The HR of the NA was 0.47 (95% CI, 0.17–1.34; *p* = 0.16). Among the 368 patients who were under observation beyond 2 years after enrollment, 70 patients started NA treatment. 45 patients developed HCC (5 patients with NAs and 40 without NAs) (Figure 4B). The HR of the NA was 0.43 (95% CI, 0.16–1.11; *p* = 0.08). To conclude, NA treatment did not significantly reduce the risk of HCC.

**Figure 4.**
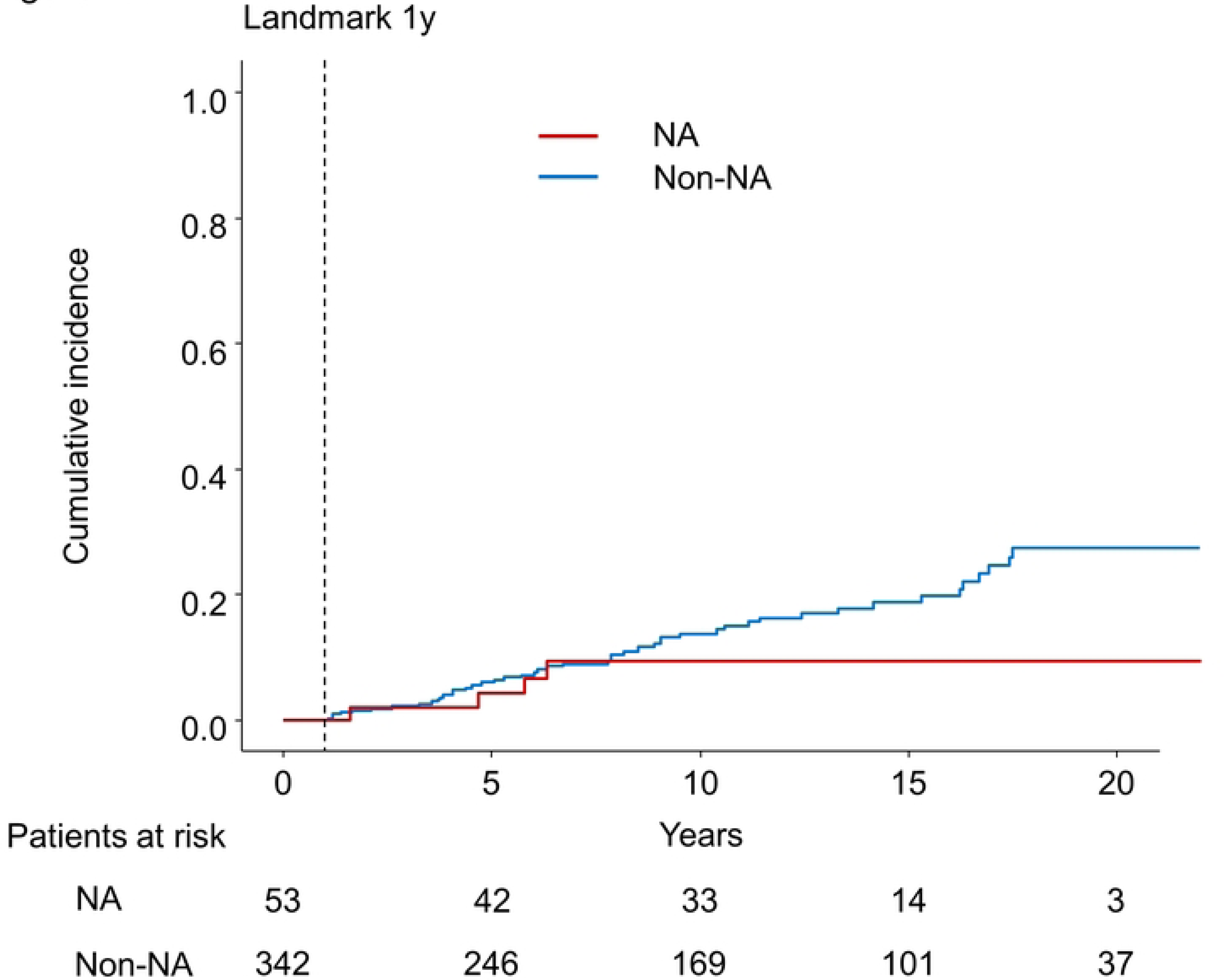
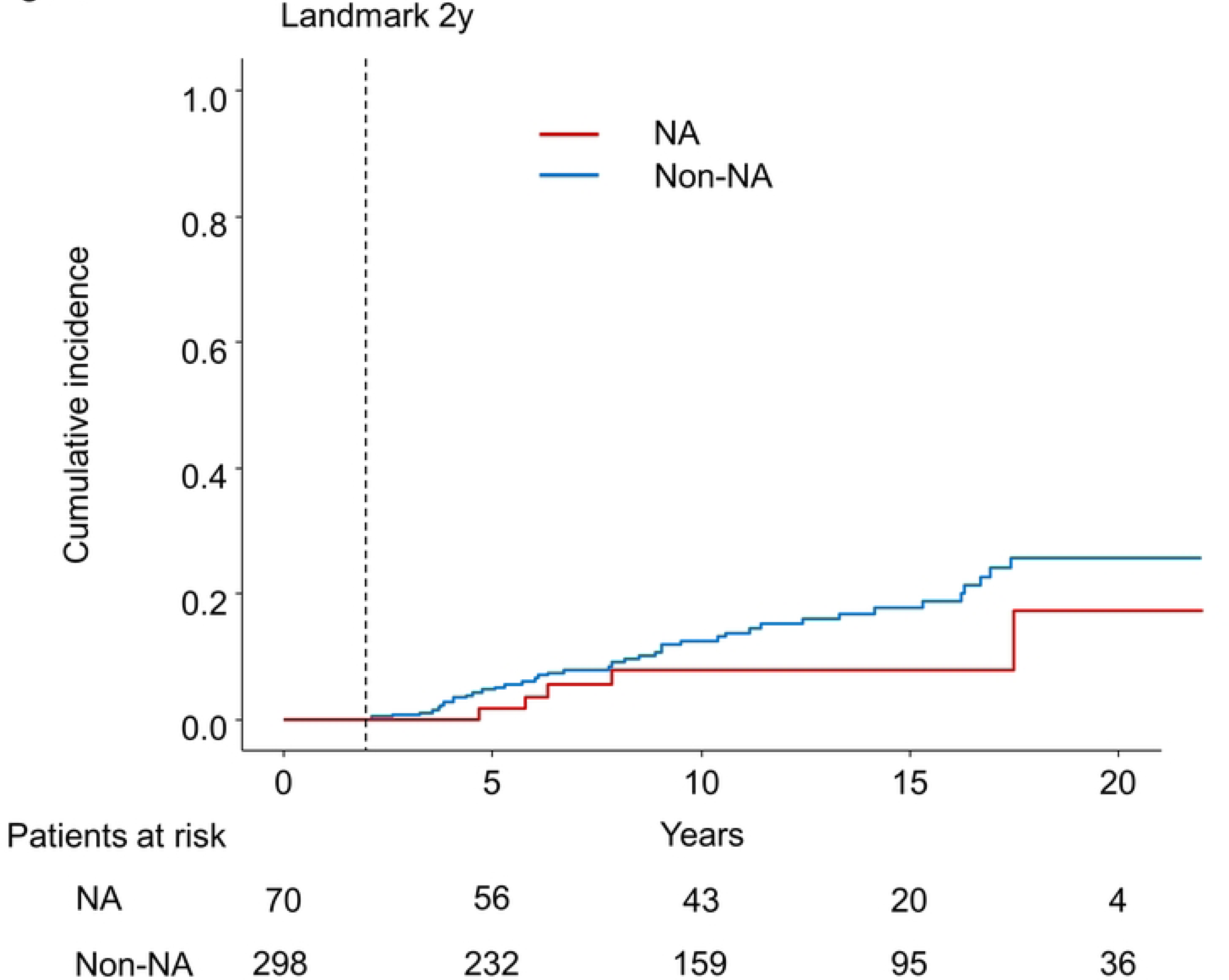
Kaplan–Meier estimate of cumulative incidence of HCC. (A) 1-year landmark analysis (B) 2-year landmark analysis NA: nucleos(t)ide analog

## DISCUSSION

To the best of our knowledge, this is the first study to evaluate the effect of NA on hepatocarcinogenesis in CHB patients using the time-dependent Cox regression analysis. We also performed 1-year and 2-year landmark analyses. Both time-dependent covariate analysis and landmark analyses showed that NA use had no significant effect on the incidence of HCC. Several observational studies have reported carcinogenic inhibitory effects of NA.[15–20] A cohort study with PSM by clinical background showed that ETV use reduced the risk ratio of carcinogenesis to 0.37.[15] A retrospective cohort study of 1870 patients reported a lower 5-year carcinogenesis in patients with cirrhosis, with a 0.55 risk ratio in the ETV-treated group compared to the historical control group.[19] One possible reason for inconsistent results with previous reports regarding the suppressive effect of NA on hepatocarcinogenesis might be the selection bias: those expected to have a worse prognosis tended to be initiated on NA therapy. NA use was paradoxically correlated with increased risk of HCC in the univariable analysis, which became insignificant in the multivariable analysis using other significant risk factors.

The adjustment for confounding factors might not be enough to see a tumor preventive effect of NA or the reported effect of NA on HCC incidence might be overestimated. Indeed, the incidence of hepatitis B-associated HCC has not changed over the past two decades, despite NA being widely available in Japan.[29]

Another possible explanation for the insignificant results is that the effect of NA was not sustainable due to the presence of resistant strains. Some reports indicated that entecavir, which yields higher potency for viral suppression than lamivudine, could suppress the HCC incidence more effectively.[15, 30, 31] However, in more than 70% of the patients started NA treatment with ETV, TDF, or TAF, the treatment response was stable and sustained. In addition, most of the patients receiving LAM were treated with ADV add-on at viral breakthrough and finally switched to ETV, TDF or TAF. As a result, most patients receiving NAs achieved SR at the end of the follow-up period, suggesting a sufficient anti-viral effect of NAs.

It is also known that it takes several months to a year to achieve SR with NAs.

Therefore, it might take a certain period to see the anti-carcinogenic effect of NAs after the initiation of treatment. However, the cumulative incidence curve plotted according to the Simon and Makuch method did not show a delayed effect of NAs. The landmark analysis also did not show the impact.

Various factors are reported to contribute to HBV-induced carcinogenesis, such as accumulation of genetic abnormalities in the process of repeated hepatocyte necrosis and regeneration due to chronic inflammation, the induction of mutations and increased genomic instability due to incorporation of HBV DNA into the host genome, and direct action of HBV X protein, which is expressed by HBV. [32–35] NA can suppress chronic inflammation by regulating HBV replication; however, it may not suppress the integration of HBV DNA into the host genome or the direct action of HBV X protein.

Therefore, it may be difficult to completely inhibit carcinogenesis with NA.

In papers with subgroup analyses by presence or absence of cirrhosis, NA therapy reduced the incidence of hepatocellular carcinoma only in patients with cirrhosis, except for one paper [36]. Hosaka et al. reported that HCC incidence was decreased in ETV-treated patients compared to untreated patient in cirrhosis (*p* < 0.01), whereas no significant difference was observed in noncirrhosis (*p*= 0.44) [15]. In a nationwide cohort study conducted in Taiwan, Wu et al. reported NA therapy was associated with decreased HCC incidence.[20] However, the impact was smaller in noncirrhosis than cirrhosis (HR, 0.72 vs. 0.27). Our subgroup analysis also showed that NA therapy reduced the HCC incidence only in cirrhotic patients. It may take a longer duration to observe decreased HCC incidence in noncirrhotic patients.

Although the incidence of HCC did not differ between the two groups, HCC was diagnosed at an earlier stage in the NA group, which might suggest the lead-time bias. However, the lead time was not long, considering the difference in mean tumor size was only 2 mm. Another explanation is that the tumor doubling time was longer in the NA group with reduced signals of tumor progression propagated by necroinflammation, supported by the lower AST and ALT.[37]

There were some limitations to this study. First, although we performed PSM, we could not completely exclude selection bias, as the use of NA is recommended for those at higher risk of HCC according to clinical practice guidelines. Second, because the unit of DNA differs depending on the time of measurement, we had to set a single cut-off point for HBV DNA by converting different units according to the conversion formula. More precise analysis using continuous variables may yield a different result.

In conclusion, our study did not reveal a preventive effect of NAs on hepatocarcinogenesis in patients with CHB.

## Data Availability

All relevant data are within the paper and its Supporting information files.

## Acknowledgments

We would like to thank Editage (www.editage.com) for English language editing.

## Author contributions

Conception and design: Makoto Moriyama, Ryosuke Tateishi.

Acquisition of the data: Makoto Moriyama, Ryosuke Tateishi, Mizuki Nishibatake Kinoshita, Tsuyoshi Fukumoto, Tomoharu Yamada, Taijiro Wake, Ryo Nakagomi, Takuma Nakatsuka, Tatsuya Minami, Masaya Sato

Analysis and interpretation of the data: Makoto Moriyama, Ryosuke Tateishi. Drafting of the manuscript: Makoto Moriyama, Ryosuke Tateishi.

Statistical analysis: Makoto Moriyama, Ryosuke Tateishi.

Study supervision: Ryosuke Tateishi, Mitsuhiro Fujishiro, Kazuhiko Koike. Final approval: All of the authors.

Agreement to be accountable for all aspects of the work: All of the authors.

## Conflict of interest

Kazuhiko Koike has received research funding from Bristol-Meyers Squibb, Gilead Sciences.

Ryosuke Tateishi has received lecture fee from Bristol-Meyers Squibb, Gilead Sciences. Tatsuya Minami has received lecture fee from Gilead Sciences.

## Abbreviations

ADV, Adefovir; AFP, alpha-fetoprotein; AFP-L3, lens culinaris agglutinin-reactive fraction of alpha-fetoprotein; ALT, alanine aminotransferase; AST, aspartate aminotransferase; CHB, chronic hepatitis B; CI, confidence interval; CT, computed tomography; DCP, des-gamma-carboxy prothrombin; ETV, Entecavir; HBV, hepatitis B virus; HBeAg, hepatitis B e antigen; HBsAg, hepatitis B surface antigen; HCC, hepatocellular carcinoma; HR, hazard ratio; LAM, Lamivudine; MRI, magnetic resonance imaging; NA, nucleos(t)ide analog; PLT, platelet count; PCR, polymerase chain reaction; PSM, propensity score matching; SR, sustained virological response; TAF, Tenofovir alafenamide; TDF, Tenofovir disoproxil fumarate

## Financial Support

Japan Agency for Medical Research and Development (AMED): JP22fk0210066 and JP22fk0210090; The Health, Labour, and Welfare Policy Research Grants from the Ministry of Health, Labour, and Welfare of Japan: H30-Kansei- Shitei-003.

---

Supplementary Figure 1. Schematic presentation of the immortal time bias

Scenario 1: Patients are divided according to the use of nucleos(t)ide analogs (NAs) at baseline. Patients 3 and 4 may have benefited from NAs even though they are categorized in the non-NA group.

Scenario 2: Patients are divided according to the use of NAs during the observation period. There is the immortal bias: Patients 3 and 4 are guaranteed to be HCC-free up to the point of NA initiation.

Supplementary Figure 2. Nucleic acid analogs and viral response.

(A) Viral response in patients initiated on lamivudine. (B) Viral response in patients initiated on entecavir, tenofovir disoproxil fumarate, and tenofovir alafenamide. LAM, Lamivudine; ADV, Adefovir; ETV, Entecavir; TDF, Tenofovir disoproxil fumarate; TAF, Tenofovir alafenamide

Supplementary Figure 3. Kaplan-Meier curve for overall survival after HCC development. HCC, hepatocellular carcinoma; NA: nucleos(t)ide analog

